# Contrasting Approaches in Advance Care Planning in Dementia as Perceived by General Practitioners

**DOI:** 10.1101/2025.06.06.25329142

**Authors:** Jenny T van der Steen, Xu Jingyuan, Willemijn Tros, Jeanet W Blom

## Abstract

**Background:** Various approaches to advance care planning are being used, also for persons with dementia and their care partners. Two contrasting approaches involve a highly scripted, predominantly medical approach to decide on specific treatments in advance versus a more flexible psychosocial, coping-based approach comprising global care goal setting.

**Objective:** To assess situations in which either approach is preferred in dementia from the perspective of general practitioners.

**Methods:** We interviewed thirteen practitioners participating in the Dutch CONT-END study; seven were trained in the medical approach and six were trained in the psychosocial approach. We explained the other approach during the interview. Twelve other practitioners were interviewed after viewing video vignettes of the two approaches shown in random order. Inductive qualitative content analyses was guided by the aim to elucidate for whom and when an approach was preferred.

**Results:** Four attributes distinguished situations in which either approach is preferred: understanding, trust, readiness and momentum. For the medical approach, understanding, trust, and readiness on part of person and care partner were prerequisites for optimal momentum, which, however could also be triggered by urgent medical reasons. In contrast, the psychosocial approach would help understand the person, foster trust and create readiness from a first conversation. Without a clear trigger, however, momentum would need to be created.

**Conclusions:** Skill in employing various approaches to ACP conversations each with specific benefits could help tailor ACP to the individual and their situation. Further theoretical and empirical research including in other populations and settings may inform person-centred ACP.

**Key Points:** A medical versus a more psychosocial approach to advance care planning (ACP) in dementia can be typified by contrasts within four attributes: understanding, trust, readiness and momentum.

Although both approaches are appropriate in itself, either is superior depending on the situation of the persons involved, and the psychosocial approach may be particularly suitable for a first conversation initiated by a professional caregiver.

Further theoretical clarification and empirical research could enhance and refine ACP training programs and inform person-centred ACP in more diverse practices and populations.

## Introduction

Advance care planning (ACP) is increasingly being conceptualized as a communication process on future care which may or may not result in formal documentation [1]. However, what ACP entails exactly, has been unclear in debates about its effects [2] while there is also overlap between ACP and “goals of care discussions,” “end-of-life communication” and “future care planning” [3-5].

Some feel that ACP should result in decisions about medical treatment at the end of life. However, particularly in dementia, a broader approach addressing psychosocial or spiritual care preferences may fit as well. For example, relatedness with social rather than medical issues was strong in community-dwelling people with young-onset dementia considering a good life [6]. In line, two contrasting approaches surface from the literature, involving the highly scripted, predominantly medical approach to decide on specific treatments in advance, versus a more flexible psychosocial, coping-based approach that comprises global care goal setting.

ACP preferably starts soon after dementia diagnosis when usually the person can still be involved [7]. Therefore, general practitioners rather than palliative care or other medical specialists often bear responsibility in initiating conversations or probing readiness to ACP, or continue conversations as part of post-diagnostic support. We aimed to assess attributes that distinguish situations in which a medical or psychosocial approach is preferred from the perspective of general practitioners (GPs) as a step towards conceptual clarity regarding approaches tailored to the individual with dementia and their care partner.

## Methods

We conducted a qualitative interview study nested in a cluster-Randomized Controlled Trial (cRCT) in which we trained GPs to deliver ACP in dementia according to either the more medical and concrete, or the more psychosocial, global approach [8]. We triangulated the data with physician interview data from a vignette study explaining the two approaches [9]. In both studies, we asked interviewees to compare. Most interviewees practiced in the urbanized west of the Netherlands.

### Two CONT-END studies

In the CONT-END cRCT, we aimed to assess the effects of two ACP approaches on wellbeing of persons with dementia and on their care partners’ evaluations of decision making and communication with physicians. We collected these data between May 2021 and December 2024. GPs including nurse practitioners (NPs), and their general-practice based assistant practitioners (“POH”) were trained in small groups in either approach. They attended two 3-hour training sessions a few months apart between February 2022 and February 2024, followed by an e-learning with a digital conversation with an avatar of a person with dementia providing feedback on choices in the conversation [10]. The two in-person sessions included theory explained by a physician-trainer familiar with the respective approach, and practicing conversations with a team of professional actors who were also GPs. After the training, between February 2023 and June 2024, we invited practitioners to an interview with XJ (female psychologist, PhD candidate leading the vignette study), WT (female MD, PhD candidate) or JWB (female GP, associate professor) about their experiences with practicing the trained approach, explaining the other approach and asking to what extent either approach would fit with the person and care partner (Supplement, interview guide 1).

In the CONT-END vignette study [ref9Smaling], in the Netherlands, we recruited 50 physicians including 12 GPs to view video vignettes that explained the two ACP approaches in randomized order between July 2020 and January 2021. The study focused on acceptability of the approaches and interviews (by XJ and a senior female psychologist) contained closed and open-ended questions (Supplement, interview guide 2).

### The two approaches

The conceptualized medical approach targeted decision making, and therefore, theory was based on the revised three-talk model for shared decision making that comprises three steps: “team talk, option talk, and decision talk” [11,12]. The psychosocial approach was based on the PREPARED framework [13] “for discussing ACP and end-of-life issues with patients with advanced life-limiting illnesses and their families.” Items included, for example, “Elicit patient preferences” including their goals and values, “Acknowledge emotions and concerns” and “Realistic hope should be fostered.”

### Analyses

Inductive content analyses by JTS (CONT-END’s PI, methodologist), and XJ started with coding three cRCT interview transcripts independently. To answer the research question, we selected codes assigned for situations, i.e. for whom and when a particular approach was being preferred. We avoided assigning codes to multiple attributes, to clearly demarcate attributes and contrasts when possible. Codes were discussed and adapted by XJ and JTS after which XJ coded the remaining transcripts. Codes were then compared with the available vignette study coding as proposed by XJ, discussed and adapted by JTS.

To expose key contrasts underlying the conceptualized ACP approaches, codes preferably contained direction and meaning. Key contrasts were then placed within broad attributes that represented more general categories. We verified whether patterns of codes differed by the trained approach in the cRCT.

### Ethics

The Medical Ethics Committee-Leiden The Hague Delft CONT-END approved an amendment to the cRCT (NL71865.058.20) on 8 December 2022 to examine the two approaches in qualitative interviews rather than in underpowered quantitative analyses. They approved the vignette study (NL72354.058.19) on 10 April 2020.

## Results

We interviewed 25 practitioners in total. In the cRCT, 6 interviews were conducted with 7 practitioners trained in the medical approach (4 GPs, 1 NP and 2 assistant practitioners) and 4 interviews with 6 practitioners trained in the psychosocial approach (3 GPs, 1 NP and 2 assistant practitioners). Three were duo interviews with GP and either NP (1) or their assistant practitioner (2 interviews). Ten interviewees were female and 3 were male. Median age was 54.5 years (range: 35-57), and median experience in their function was 12.5 years (range: 1-29). The 10 practices typically (median) enrolled 2 dyads of person with dementia and care partner (1 practice enrolled none, 2 enrolled 1, 4 enrolled 2, 2 enrolled 4, and 1 enrolled 6 dyads). In the vignette study, 12 other GPs participated (6 males, 6 females). Their median age was 30 (range: 26-32), and median experience in dementia care was 2.5 (range: 1-7) years.

To 37 codes in the cRCT data, three codes unique to the vignette study were added, all referring to the medical approach. Eleven codes remained unique to the cRCT, eight of which on “readiness,” four for each approach (Supplement).

The codes linked to attributes comprised four key contrasts of the approaches: “understanding;” “readiness;” “trust;” and “momentum” (Table 1). For the medical approach, understanding, trust, and readiness were prerequisites for optimal momentum, which, could, however, be indicated for urgent medical reasons. In contrast, the psychosocial approach would help understand the person, foster trust and create readiness from a first conversation. Without a clear trigger, however, momentum would need to be created.

**Table 1.** An analysis of perceived fit of two conceptualized ACP approaches with situations (relating to the person, care partner and physician involved, and to circumstances)

| Attribute | Two contrasting approaches |  |  |  |
| --- | --- | --- | --- | --- |
|  | ACP – medical, concrete approach |  | ACP – psychosocial, global approach |  |
|  | Key contrast | Description of findings | Key contrast | Description of findings |
| Understanding | Understanding required (from person) | Understanding of the decisions at stake by the person is conditional for applying this approach or for the person to initiate a conversation, while some persons with dementia are not capable. | Understanding facilitated (for person) and offered (to physician) | Understanding of the person is offered to the physician and the physician's better understanding of the whole person, background and where they come from helps them in guiding person-centred decision making. |
| Readiness | Readiness present (person, or also care partner) | Related to understanding is the condition that the person or also the care partner are ready for ACP in a more general sense. This is most obvious through them asking for concrete treatment orders themselves or taking initiative for an ACP conversation. Direct asking was perceived as a style of communication related to personality or to regional culture. | Readiness fostered (physician, actively to person, or also care partner) | Creating or probing readiness is a mechanism of how this approach can work. In that sense, it is the most proactive approach and may be suitable before moving to the other approach for which readiness would be a requirement. |
| Trust | Trust needed | There should be trust from both sides that the relationship is not being damaged through discussing sensitive topics and the sharing of potentially confronting information. Relevance of trust is increased because families may be divided among themselves about the best treatment or they may overshadow the wishes of the person. | Trust established | A relationship of trust with the person is established or facilitated through this approach, through taking time to get to know the person in an early phase of dementia with no need for confrontational decision making about medical decisions. |
| Momentum | Momentum present (time right to express preferences) | The moment is right for the person to clarify their preferences as routine at a particular time, urgent, or a preferred moment. Caution changing preferences or scenarios not anticipated. | Momentum generated (time taken for meaning) | The moment is created when there is time to understand what is meaningful for the person. Caution topics addressed too broad and not actionable. |

Of the 40 single codes, 36 were consistently linked to a single approach, while four (for each attribute except readiness) applied to both approaches. Only for understanding there were different directions; it was undecided which approach was easier. Codes did not differ by trained approach in the cRCT.

## Discussion

A medical versus a more psychosocial approach to ACP in dementia can be typified by contrasts within four attributes: Understanding, Trust, Readiness and Momentum. Momentum could be conceptualized as situations that bring attributes and approaches together. Momentum can refer to timeliness of physician and person understanding each other, benefitting from, or creating readiness and a relationship of trust needed to engage in ACP effectively.

Both approaches are appropriate in itself as acceptable in more countries [14] and elements of both approaches were recommended in an international Delphi study [7]. However, either is superior depending on the situation of the persons involved. The psychosocial approach may be particularly suitable for a first conversation initiated by a professional caregiver who is uncertain about capacity or openness to engage in decision making or to discuss sensitive topics. In an interview study with Dutch GPs, building trust and addressing non-medical issues soon after diagnosis facilitated ACP [15]. Starting with the more scripted medical approach may fit in case of high health literacy and may be indicated regardless when health declines rapidly. Indeed, a systematic review on preferences of older people concluded that the “ACP approach should be selected carefully to match the person’s health and psychosocial status” [16].

A strength of this small study is that it was conducted in the context of a unique trial with separate arms for the two approaches. GPs were trained in one approach to be applied in practice. Despite the interviewees’ limited understanding of the other approach, the cRCT delivered more unique codes than the vignette study, in particular related to readiness (8 of 11 unique codes). Possibly actually conducting ACP triggers more thinking about the person’s readiness. Also, the GPs in the vignette study were young and less experienced. The analysts had different relatedness to the topic: JTS conceptualized approaches, XJ interviewed in both studies which helped when discussing codes.

Training skills in employing various approaches to ACP conversations each with specific benefits can help tailor ACP to the individual and their specific situation. Further theoretical and empirical research is needed including in other populations and settings to inform person-centred ACP interventions. Such interventions and its timing should effectively fit with the individual and their care partner.

## Data Availability

The codes resulting from the qualitative analyses are available upon request. There is no permission from the participants to make the interview transcripts available.

## Acknowledgement

The authors thank Dr. Marieke Perry, MD, PhD and Dr. Jentie Kraamer, MD for providing training in the two approaches, Dr. Bram Tilburgs, RN, PhD and Dr. Mandy Visser, PhD for their contributions to the cluster-Randomized Controlled Trial, Prof. Wilco P. Achterberg, MD, PhD and Dr. Hanneke Smaling, PhD for their contributions to the vignette study, and the participating practitioners for their contributions to the data collected in this study.

